# Development of a differential treatment selection model for depression on consolidated and transformed clinical trial datasets

**DOI:** 10.1101/2024.02.19.24303015

**Authors:** Kelly Perlman, Joseph Mehltretter, David Benrimoh, Caitrin Armstrong, Robert Fratila, Christina Popescu, Jingla-Fri Tunteng, Jerome Williams, Colleen Rollins, Grace Golden, Gustavo Turecki

**Affiliations:** Douglas Mental Health University Institute, Montreal, Canada; McGill University, Montreal, Canada; Aifred Health Inc., Montreal, Canada; University of Alberta, Alberta, Canada; University of Cambridge, Cambridge, United Kingdom; University of Waterloo, Waterloo, Canada

## Abstract

**Objective:** Major depressive disorder (MDD) is the leading cause of disability worldwide, yet treatment selection still proceeds via “trial and error”. Given the varied presentation of MDD and heterogeneity of treatment response, the use of machine learning to understand complex, non-linear relationships in data may be key for treatment personalization. Well-organized, structured data from clinical trials with standardized outcome measures is useful for training machine learning models; however, combining data *across* trials poses numerous challenges. There is also persistent concern that machine learning models can propagate harmful biases.

**Methods:** We have created a methodology for organizing and preprocessing depression clinical trial data such that transformed variables harmonized across disparate datasets can be used as input for feature selection. Using bayesian optimization, we identified an optimal multi-layer dense neural network which used data from 21 clinical and sociodemographic features as input in order to perform differential treatment benefit prediction.

**Results:** With this combined dataset of 5032 individuals and 6 drugs, we created a differential treatment benefit prediction model. Our model generalized well to the held-out test set and produced similar accuracy metrics in the test and validation set with an AUC of 0.7 when predicting binary remission. To address the potential for bias propagation, we used a bias testing performance metric to evaluate the model for harmful biases related to ethnicity, age, or sex.

**Conclusion:** We present a full pipeline from data preprocessing to model validation that was employed to create the first differential treatment benefit prediction model for MDD containing 6 treatment options.

## Introduction

The World Health Organization estimates that major depressive disorder (MDD) impacts more than 300 million people worldwide (World Health Organization, 2017). Despite the availability of several effective depression treatments, many patients undergo the inefficient “trial and error” approach to treatment selection, which can result in lost time and worse disease outcomes (Kraus et al., 2019). Considering the heterogeneity of depression and treatment response, it would be of significant value to identify the optimal treatment specific to a particular patient’s characteristics (Benrimoh et al., 2018; Mehltretter et al., 2020).

Machine learning (ML) methods are well-suited for the challenge of developing personalized treatment approaches in psychiatry (Benrimoh et al., 2018; Mehltretter et al., 2020). Moreover, deep learning, a ML technique that uses artificial neural networks to learn high-level representations from raw data and model complex relationships between variables, is particularly suited for predicting depression treatment response and facilitating effective personalized treatment (see review of previous work in (Durstewitz et al., 2019; Squarcina et al., 2021).

A recent review summarizing 8 studies using deep learning methods to predict treatment response in depression found that models generally reached AUCs (area under the curve, a threshold-free measure of a model’s ability to classify successfully) between 0.66 and 0.82 (Squarcina et al., 2021). While some of these studies incorporated functional and structural magnetic resonance imaging, genetics, and epigenetics as model features, it is not currently feasible to collect biomarker data in routine practice which leaves symptom/clinical and sociodemographic features as the most practical potential predictors (Perlman et al., 2019). One key limitation of most of the studies reviewed is that they aimed to predict treatment response between two treatments or of one treatment at a time, whereas the clinical reality that clinicians and patients face is selecting a treatment from many options: over 20 antidepressants, psychological therapies, and neuromodulation treatments (Cipriani et al., 2018).

Another limitation of machine learning models is the potential for propagation or amplification of harmful biases (Mehltretter et al., 2020; Tanguay-Sela et al., 2022). For example, it is crucial to be conscious of, and address, the possibility that a model that learns to predict worse remission rates for patients from a particular background than what is actually observed for those patients in the data (assuming that the data is reasonably representative of the intended use population, which may not always be the case).

In previous work, we demonstrated the use of a neural network capable of differential treatment benefit prediction (that is, the generation of remission probabilities) for a number of treatments (Mehltretter et al., 2020). In this work, we expand on our previous work in two key ways. The first is by addressing the problem of dataset merging. Datasets must be merged for two reasons: to generate a sufficient sample size for model training, and to provide the model with examples of patients on a number of different treatments in order for the model to be able to learn to differentiate between treatments. When working with data from a number of different sources, a significant challenge is the heterogeneity of study design and data collection: different studies used a variety of scales and questionnaires to assess depression symptoms and other psychopathology, physical health, and well-being outcomes (i.e.., comorbid psychiatric symptoms and quality of life). Driven by shortcomings in the clinical utility of diagnostic categories and the phenomenological overlap of symptoms and traits across diagnoses, Waszczuk et al., produced a hierarchical system focused on the dimensionality of emotional disorders (e.g., somatoform, internalizing, detachment) and the various manifestations of these disorders that fall within each category (Waszczuk et al., 2017; Ruggero et al., 2019). For example, OCD is classified under the “fear” category, which itself is classified under the “internalizing” category (Ruggero et al., 2019). While HiTOP shows potential advantages in both clinical utility and research,, this taxonomy structure is limited to psychopathology dimensions; it does not represent other patient-level characteristics that are vital for predicting treatment response, such as demographic information (e.g., years of education, socioeconomic status), personal history (e.g., trauma), or physical health (e.g., body weight (Puzhko et al., 2020), comorbid health problems) (Perlman et al., 2019). It similarly does not capture the health outcomes necessary for understanding a patient’s well-being and response/remission to treatment, such as daily functioning and quality of life. Here we present a detailed methodology for a novel taxonomy (inspired by the HiTOP method) and variable transformation procedure that was created in order to facilitate data collation and model training.

The second expansion of our previous work is to include a more robust assessment of learned model bias, in order to ensure that models put into clinical practice do not propagate harmful biases. Below, we present our differential treatment benefit prediction model results, subgroup analyses arising from our bias testing, and the patient features retained by the model.

## Methods

The end goal of our work was to produce a model capable of predicting remission and generating differential treatment benefit predictions for a number of different treatments. We specifically selected remission, based on a cutoff of 10 points on the MADRS and 7 points on the HAM-D, as the main outcome measure because it is both binary and the gold-standard objective in depression treatment (Kennedy et al., 2016).

### Data

De-identified patient-level data from clinical trials of depression treatment were provided by GlaxoSmithKline and Eli Lilly via the Clinical Study Data Request (CSDR) platform, along with the relevant study protocols. These data sets were chosen because of their accessibility in a digitized format and their heterogeneity compared to the data used in our previous work. Our inclusion criteria for data were simply that the primary indication was depression (comorbidities are allowed) and that outcomes were measured using standardized rating scales. Studies were excluded if they measured a pediatric population, if the patients had bipolar disorder, or if their depression was caused by substances or by another medical condition. After the study selection process illustrated in Figure 1, we were left with 21 included studies. Supplementary Table 1 shows the breakdown of the studies included and the sample size (n) of the patients given each medication in each study.

**Figure 1.**
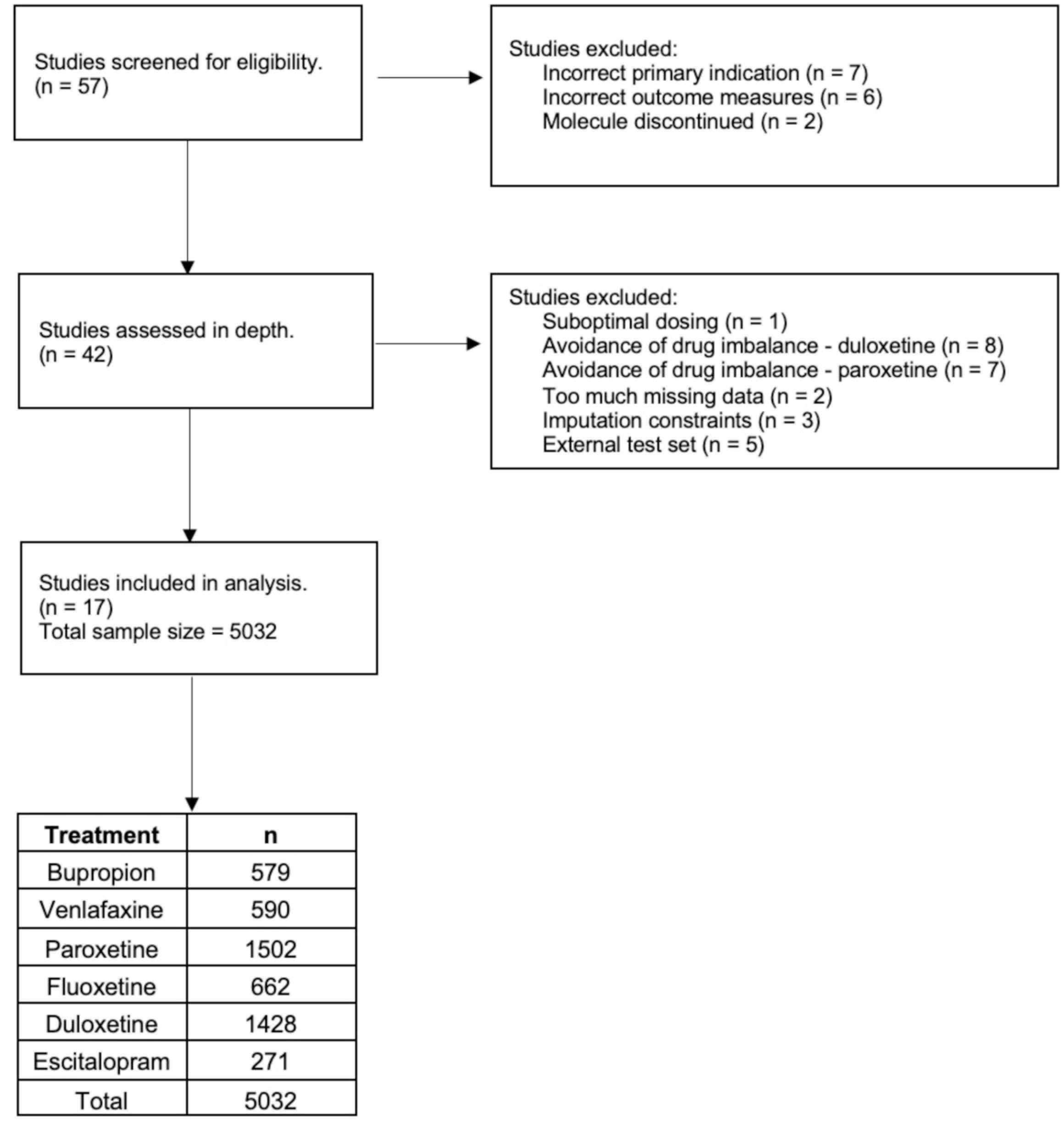
Flowchart summarizing study selection. Adapted PRISMA diagram indicating how individual studies were reviewed and selected for analysis, along with reasons for exclusion. “Avoidance of drug imbalance” refers to withholding studies to avoid having an overrepresentation of a given drug, reducing the risk of potential bias.

### Data Preprocessing and Taxonomization

The main steps taken to preprocess and prepare the data for merging are summarized in Figure 2. We began by extracting and standardizing all the questionnaires from the datasets corresponding to these studies. This involved creating standard question texts and response values for each individual question of each questionnaire. This resulted in 57 standard questionnaires. According to the associated study protocols, as well as the raw data itself, we identified the version of the questionnaires that were used in the respective studies, in cases where several versions existed across studies (e.g., short form vs long form) or where differences in question phrasing/text were found (Fenton & McLoughlin, 2021).

**Figure 2.**
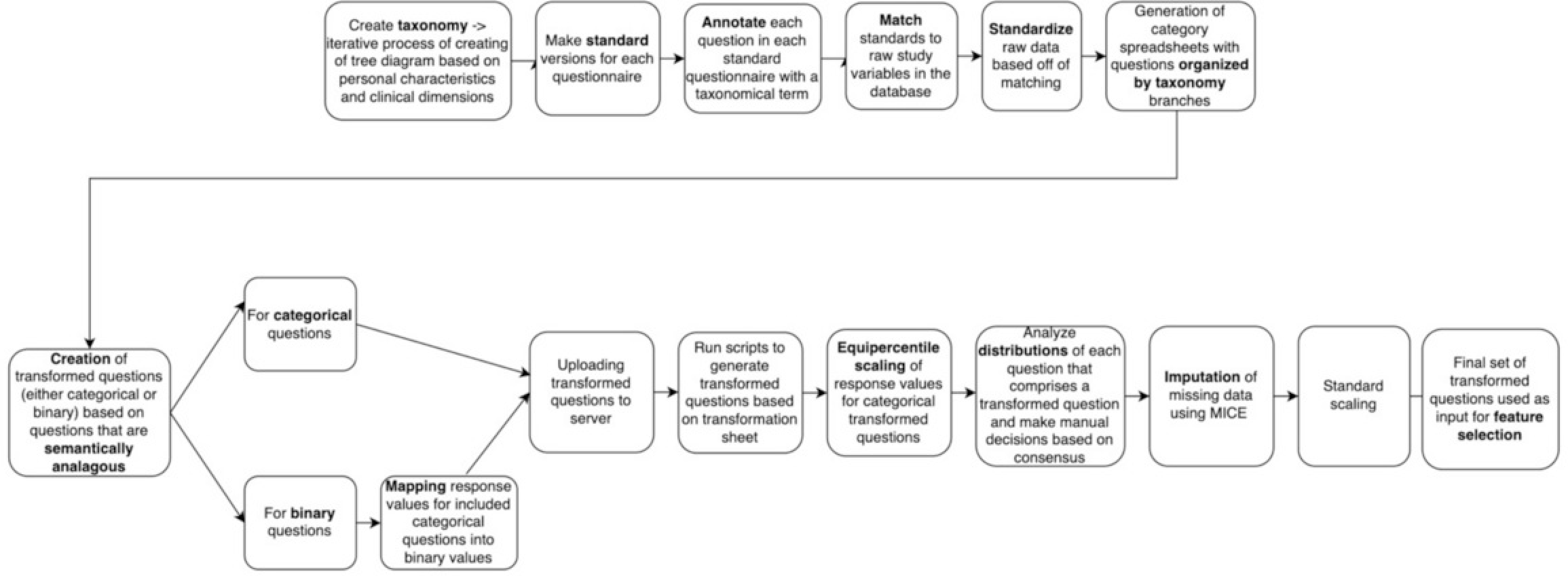
Flowchart for custom data preprocessing and transformation. The process illustrated here covers the main steps from creating a standardized classification system for questions based on a taxonomy tree all the way to having a finalized set of transformed and scaled input features to use for feature selection.

In parallel, we created a custom taxonomic system to categorize our data spanning across different clinical and demographic dimensions. The taxonomy was originally inspired by the work of Waszczuk et al. (2017). While including some of the emotional symptom-based categories defined by Waszczuk et al. (2017) examples of additional higher order clusters in our taxonomy were those defining sociodemographic, physiological, cognitive, and quality of life features. Overall, our taxonomy included 17 roots (i.e., higher order categories), each containing a number of branches and leaves. As can be seen in Figure 3, quality of life root that itself has further leaves and another sub-branch (relationships), which itself has leaves (family, social, romantic).

**Figure 3.**
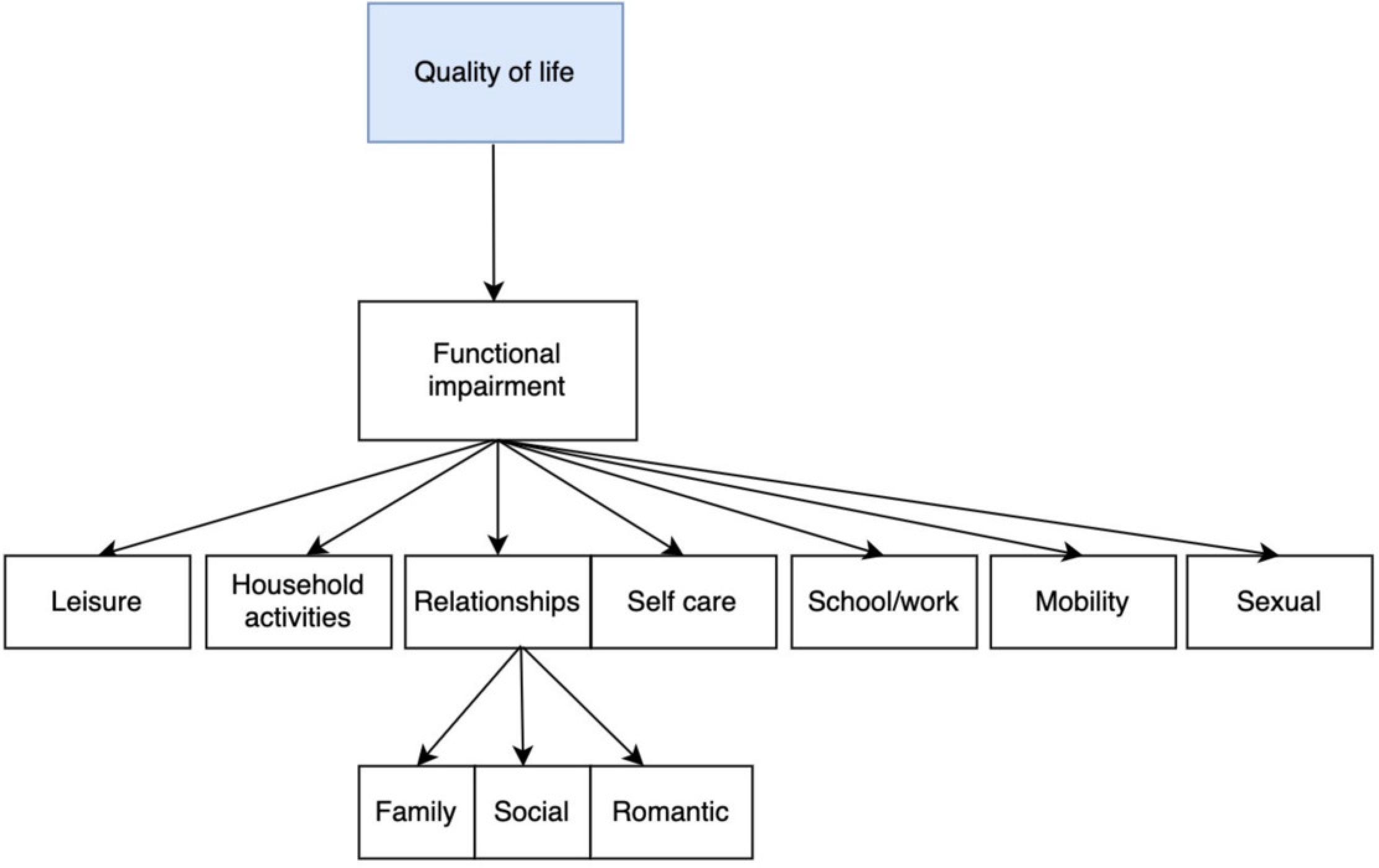
Example of taxonomical category. Functional impairment category is a branch of the quality of life root that itself has further leaves and another sub-branch (relationships), which itself has leaves (family, social, romantic).

With this taxonomy system, we tagged each individual standardized question with a root category, then used the branches to provide further categorical resolution. However, if the question was not able to be categorized using the lowest dimension it would be incorporated at the level at which it reflected the semantic meaning or clinical dimension of that feature, occasionally belonging to a higher order category as necessary. For example, for a question about functional impairment in the context of personal relationships, it cannot be classified into any of the “family”, “social”, or “romantic” leaves, as the specific relationship is not specified; therefore, the most granular category it can accurately be attributed to is “relationships”. We also allowed questions to be labeled with a secondary category in cases where this was deemed appropriate by 2 raters. Finally, we created flags that represent a certain characteristic of a question (e.g., whether it was specifically patient or clinician rated, or referring to a past time point, etc…). See Supplementary Table 2 for a glossary of terms involved in the data pre-processing and taxonomy.

During this exercise, the categories assigned to each question were assessed by at least 2 raters. Disagreements were resolved by discussion and group consensus, which ensured there was reliability and consistency of categorization. With the questions sorted by category, we created “transformed questions” that would allow for semantically similar or identical standard questions to be combined into the same feature, which could then serve as input to the predictive model. We then matched the raw data to these standards and inserted into the standardized database, generating a unique identifier for each standardized question.

This taxonomical organization helped to group all matched questions that belonged to the same high-level category. We could thus visualize all the questions within this high-level category and combine questions according to their lower-level category when appropriate. For example, if a question asked about overall functional impairment resulting depression, it would be tagged in the quality of life → functional impairment category, but if a question asked specifically about functional impairment as it relates to ability to care for oneself, it would be tagged in the quality of life → functional impairment → self-care category (Figure 3).

While attempts were made to produce features at the lowest level of categorization (i.e., the most granular level), we did not combine questions where semantic differences existed and where increased resolution would come at the expense of feature validity as assessed by raters and group discussion. Rather, we went back to the next highest level of the tree to encompass the questions at the more granular level, when this was appropriate (specifically, avoid over grouping items in any way that comprises feature validity). One priority was to combine questions that had categorical or continuous, rather than binary, response values, in order to minimize the loss of resolution which would occur when a continuous-response question was binarized. Questions that were to be combined were first rescaled by equating the smaller scale variables to the largest scale variable. In doing so all variables that could be combined based on semantic similarity were all on the same scale. The specific equating method used was dependent on the value distribution of the variables to be combined. We iteratively compared each grouped variable with the max-scale variable and if both variables to be equated had a normal distribution then linear equating was the method used; otherwise, equipercentile equating was used (Kolen & Brennan, 2014). Once questions within a category were all on a similar scale, we created our new transformed variable by taking the average value of all the variables for each patient. During scaling and transformation of categorical variables, as a validity check, we confirmed that the variance among the categorized values was not larger than 1; this value was chosen because for the type of variables we were merging, a change in value of 1 indicates a new level of severity (i.e., a change from ‘often’ to ‘very often’ on a question about a given symptom). Additionally, we confirmed using bar plots that the distribution of the transformed variable did not significantly vary from the variables that were averaged. By testing the variance of grouped variables after rescaling we could identify if any single variable within a group did not belong. Once equating was verified, we scaled all continuous variables using a standard scaler and added the minimum value plus a constant of 0.01 to variables where a value of less than or zero occurred. This additional constant was added due to how some neural network activation functions treat negative and zero values irregularly (Lederer, 2021).

If all questions within a feature were binary, no transformation was necessary. In cases where a mixture of binary and categorical questions existed within a particular semantic category, and there was a preponderance of binary questions, the entire semantic category was “binarized", meaning that each response value was given a value of either 0 or 1, depending on the response value text and how it relates to the transformed question semantics. As a validity check, we verified that the binarized version of the categorical questions followed similar response distributions as the native binary questions in the same category. That way, we could be more confident that our binarization procedure did not introduce artifactual variability. Any conflict was settled by group consensus. Moreover, having a binary cutoff on a categorical question allowed for thresholding by symptom dimension intensity when necessary. The response value transformations for all of the included transformed questions can be found in Supplementary file 1 (spreadsheet). In summary, with all the questions in our dataset matched to standards, and these standards taxonomized, we could group semantically similar questions - those querying the same dimension - into a common “feature”.

### Feature Selection

There exist many methods for feature selection, many of which rely on tree, linear or logistic regression algorithms. However, in some situations the optimal feature set selected is only optimal when used by the underlying algorithm. As we are using neural networks for our final classification model (based on superior performance of these model types in previous work (Mehltretter et al., 2020), we decided to use neural networks for our feature selection task in the form of a new layer called CancelOut *(Borisov et al., 2019).* CancelOut is a fully connected layer allowing us to create a classification model with the same task of training with a target of remission. The CancelOut layer has a custom loss function that works as a scoring method so that by the end of training we can view and select features based on their score.

We first trained one neural network with the same specifications as the network we were testing with the additional CancelOut Layer as the first layer. Because this layer cannot be used in the final neural network model that will be used for testing and inference in the clinic, as this model must necessarily input only the chosen features to reduce the burden of data collection on patients and clinicians. The number of features retained was a feature fed into our bayesian optimization framework (see below) that was then used for choosing the optimal set of hyperparameters.

### Performance metrics

Our model selection process using Bayesian optimization focused on optimizing the highest Area Under the Receiver Operating Curve (AUROC, often abbreviated as AUC), as it allowed us to understand if we were able to well separate between patients expected to remit or not remit. Since this metric is scale-invariant (i.e. more focused on the ranking than on the prediction of absolute values) and classification-threshold-invariant (focused on the effectiveness of the predictions irrespective of where the classification threshold is set), we can get a holistic and well-rounded view on the quality of the model and its performance (Jin Huang & Ling, 2005). Finally, Positive Predictive Value (PPV), Negative Predictive Value (NPV), Sensitivity and Specificity are also used as these are commonly used in clinical studies and are interpretable to both machine learning engineers and clinicians, and are clinically relevant. Multi-component metrics <u>such as these</u> (e.g., PPV + NPV and Sensitivity + Specificity) provide additional granularity on the model’s ability to identify the positive and negative class independently. This granularity can aid in determining the clinical tolerance for false positive and false negative samples so that the model can be tailored correctly. All of these metrics leverage True Positives (TP), True Negatives (TN), False Positives (FP), and False Negatives (FN). The PPV is calculated as TP / (TP + FP) and the PPV as TN/(TN+FN). The sensitivity is calculated as TP/(TP+FN) and the specificity as TN/(TN+FP) (Tharwat, 2020).

Sensitivity analyses were performed wherein we systematically varied the inputs of just one variable while holding all other variables constant (Engelbrecht et al., 1995). Examining the resulting impact on model output predictions is useful for examining the directionality and impact of individual variables on predictions and ensuring that these cohere with established literature.

### Bias Testing

Finally, we consider bias testing as an important performance metric. We therefore compare the actual population remission rate to the average predicted remission rate for each of our accessible demographic factors (age, race, and sex) and confirm that there is not more than a 5% under-prediction of remission rates and produce subgroup analyses which examine the performance of the model on subgroups in different data splits.

## Results

### Selected Studies and Final Dataset

A total of 57 studies were screened for inclusion in our analysis (Figure 1). Of these, 16 were excluded on the basis of having either an incorrect primary indication (N = 7) or outcome measure (N = 6) or having involved the study of a drug that has since been discontinued (N = 2). Of the remaining 42 studies, 21 were excluded on the basis of demonstrating suboptimal dosing regimens (Kennedy et al., 2016) (N = 1), and to avoid overrepresentation of certain medications for which large amounts of data were available (Duloxetine, Paroxetine, N = 15). A further five were removed as a result of too much missing data. A total of 21 studies remained, containing data on 5032 patients and six treatments/medications, with absolute data point numbers ranging from 271 (escitalopram) to 1502 (paroxetine). Sociodemographic data (age, sex, race/ethnicity) for the population can be found in Table l. These studies were then processed according to the procedures outlined in the methods section of this paper (Figure 2). The dataset splits had the following sample sizes: train = 4099, validation =, 422, test = 511. In total, the 21 studies contained data on six treatments/medications, with absolute data point numbers ranging from 271 (escitalopram) to 1502 (paroxetine). These studies were then processed according to the procedures outlined in the methods section of this paper (Figure 2). The treatments covered three major drug classes: selective serotonin reuptake inhibitors (SSRI), serotonin and norepinephrine reuptake inhibitors (SNRI), and a norepinephrine and dopamine reuptake inhibitor (NDRI). Supplementary Table 1 shows the drug breakdown of the 21 studies included. The real population remission rate prior to modelling was 43.18%.

**Table 1.**
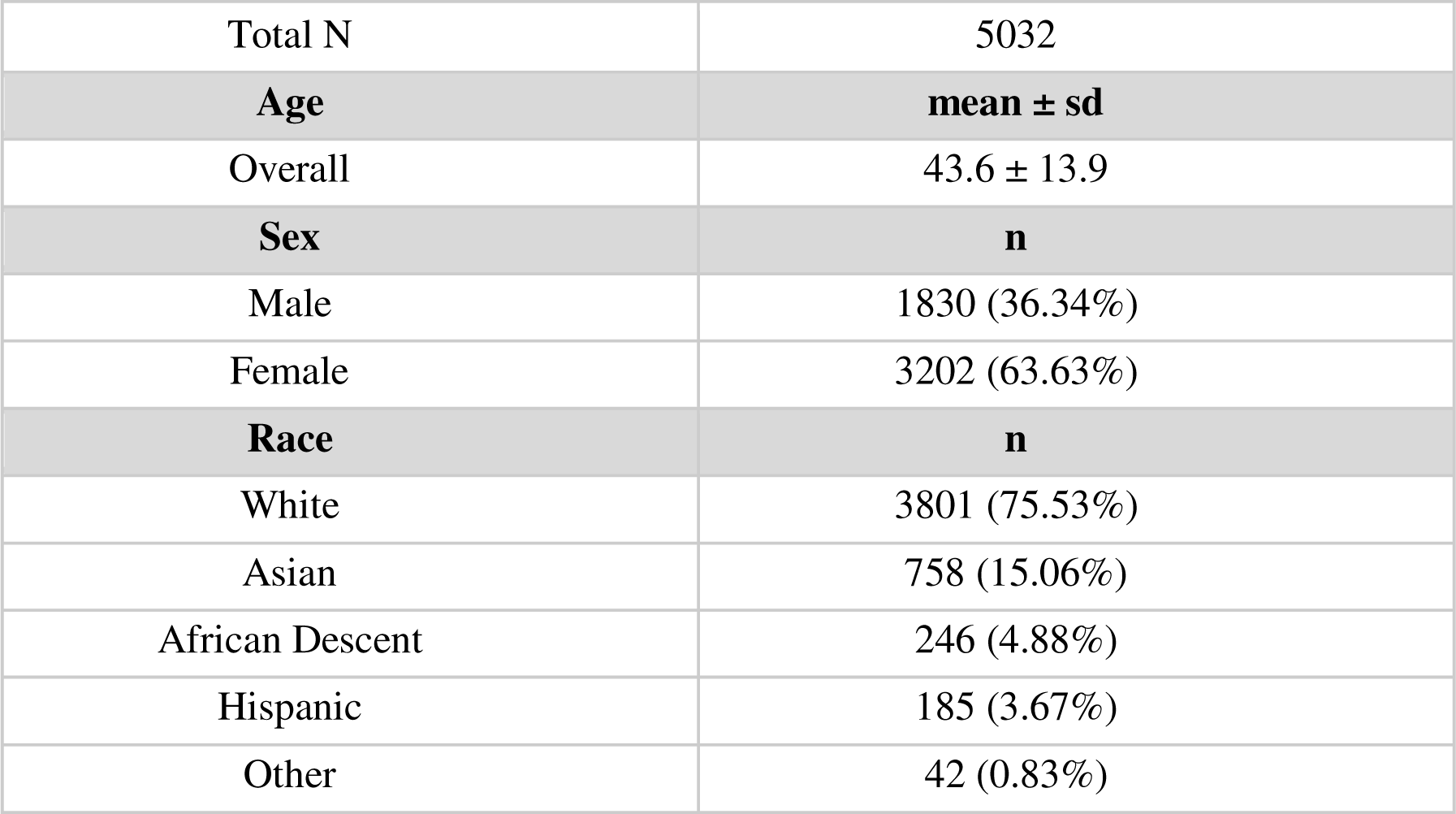
Sociodemographic data for all patients across datasets.

|  |  |
| --- | --- |
| Total N | 5032 |
| **Age** | **mean ± sd** |
| Overall | 43.6 ± 13.9 |
| **Sex** | **n** |
| Male | 1830 (36.34%) |
| Female | 3202 (63.63%) |
| **Race** | **n** |
| White | 3801 (75.53%) |
| Asian | 758 (15.06%) |
| African Descent | 246 (4.88%) |
| Hispanic | 185 (3.67%) |
| Other | 42 (0.83%) |

To identify the optimal structure for our dense neural network (DNN) we employed bayesian optimization with our package, Vulcan. Bayesian optimization allowed us to test various network configuration and features sets to test for optimality based on a set accuracy metric. For our testing purposes we want to find a DNN structure and feature set that maximized the AUC value. Our bayesian optimization testing produced an optimal model that had two-hidden layers both with 40 nodes that used exponential linear units (ELU) and a dropout value of 0.15 to assist with generalization (Clevert et al., 2015). The prediction layer determined our remission probabilities through use of the softmax function. During training the network parameters were tuned using the Adam optimization algorithm with a learning rate of 0.001. This architecture was trained with early stopping to prevent the network from overfitting. Specifically, we had 300 epochs set as the maximum with an early stopping patience of 100, which resulted in the model using all 300 epochs to train. Table 3 demonstrates the basic statistical metrics used to analyze our data, stratified by the 3 respective test sets used.

### Model Performance

Table 2 lists the 26 features (both categorial and binary), including those developed with our custom taxonomy and transformation process, which were included in our final model. Please see Supplementary Figure 1 for a visual representation of missingness score per feature by study. We achieved accuracies of 65-66% on all data splits and AUCs of 0.65-0.7 (Table 3), results which are in line with previous work (see Chekroud et al., 2016; Mehltretter et al., 2019) despite the inclusion of a larger number of medications and the merging of several datasets. We maintained reasonable F1 scores, indicating that the model achieved a balance of precision and recall. For comparison, a logistic regression model was run using the same features and data splits; it achieved an AUC of 0.62 on the test set, underperforming the neural network model (see Supplementary materials for results). It is important to note that these predictors for the logistic regression were selected by the deep learning model as it trained which is in line with common practices of using the same model for feature selection and model training.

**Table 2.**
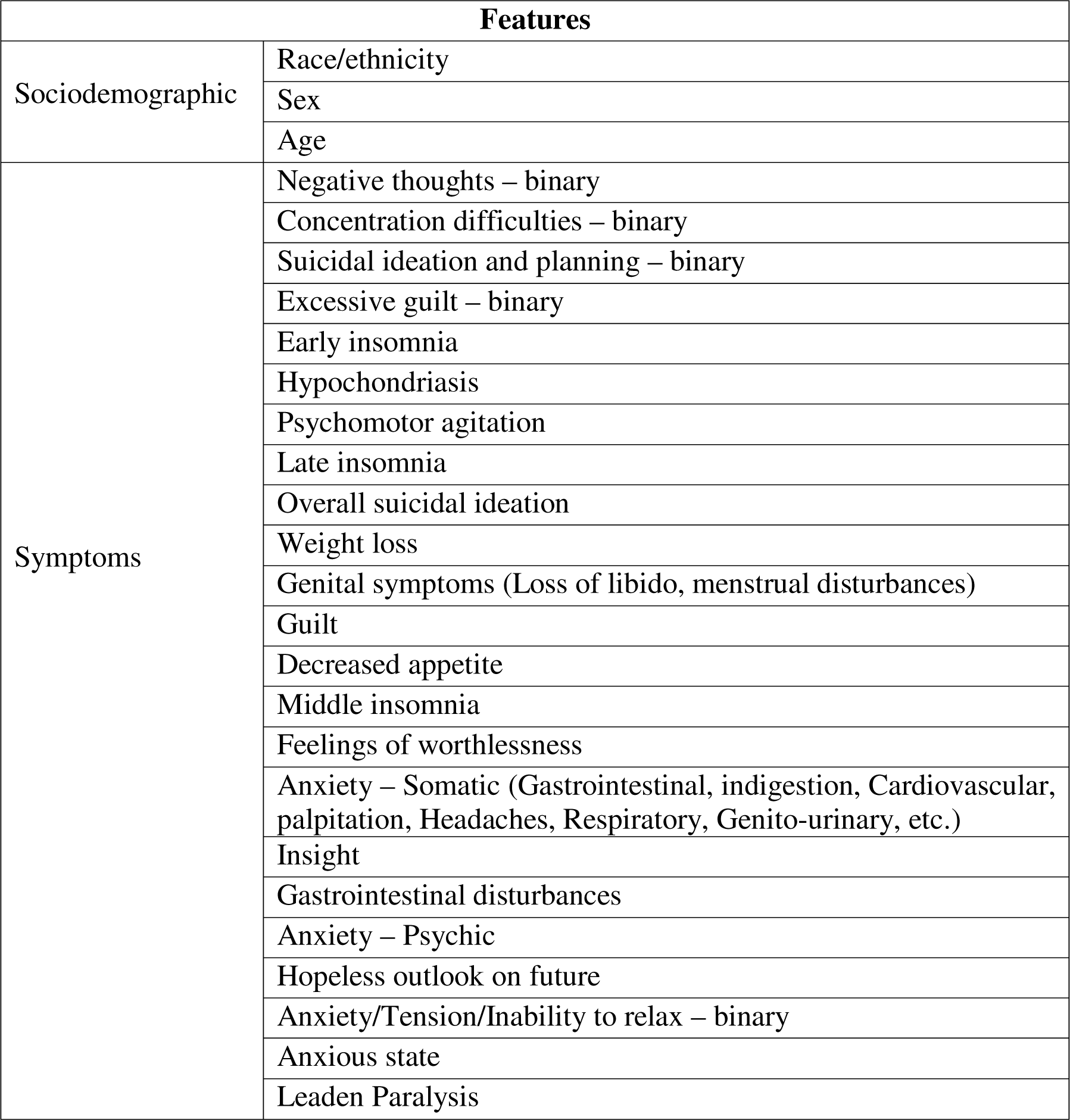
The feature table depicts the top 26 features included in the model.

| Features |  |
| --- | --- |
| Sociodemographic | Race/ethnicity |
|  | Sex |
|  | Age |
| Symptoms | Negative thoughts – binary |
|  | Concentration difficulties – binary |
|  | Suicidal ideation and planning – binary |
|  | Excessive guilt – binary |
|  | Early insomnia |
|  | Hypochondriasis |
|  | Psychomotor agitation |
|  | Late insomnia |
|  | Overall suicidal ideation |
|  | Weight loss |
|  | Genital symptoms (Loss of libido, menstrual disturbances) |
|  | Guilt |
|  | Decreased appetite |
|  | Middle insomnia |
|  | Feelings of worthlessness |
|  | Anxiety – Somatic (Gastrointestinal, indigestion, Cardiovascular, palpitation, Headaches, Respiratory, Genito-urinary, etc.) |
|  | Insight |
|  | Gastrointestinal disturbances |
|  | Anxiety – Psychic |
|  | Hopeless outlook on future |
|  | Anxiety/Tension/Inability to relax – binary |
|  | Anxious state |
|  | Leadens Paralysis |

**Table 3.**
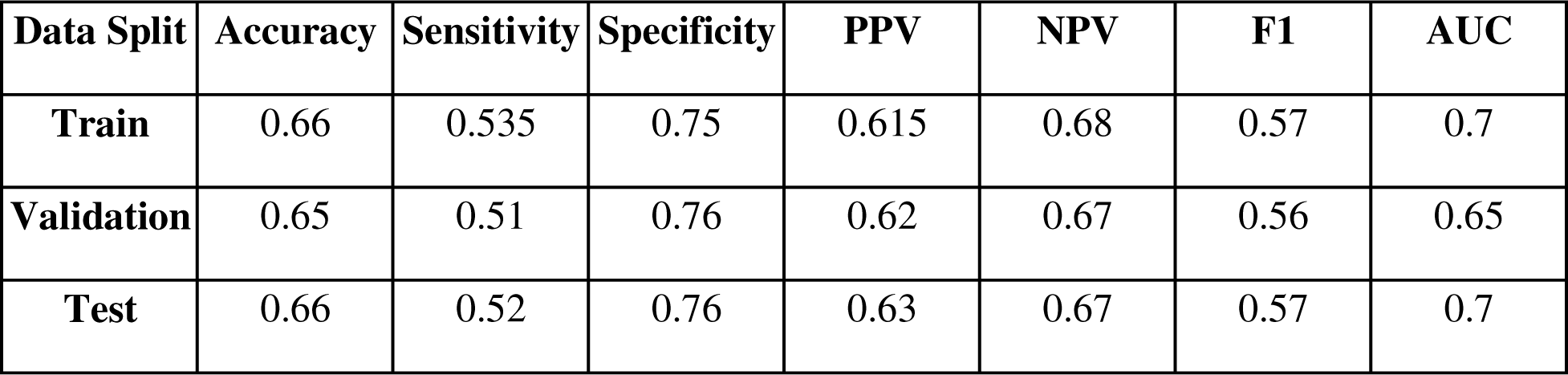
Demonstration of basic statistical metrics, stratified by the 3 sets of data used.

Examples of sensitivity analysis results are presented in Figure 4. We found that these cohered with previous literature and generally with clinical experience (Perlman et al., 2019). For example, the proportion of patients remitting decreased as we increased the suicidal ideation score; this coheres with a systematic metareview finding that suicidality was a predictor of treatment response (Perlman et al., 2019). Similarly, as concentration difficulty and leaden paralysis scores increase (indicating worsening symptomatology) the proportion of patients remitting decreases. We note the cognitive impairment, related to poor concentration, has been found to be predictive of reduced response to antidepressants, and that psychomotor retardation has been linked with worse response to SSRIs (Perlman et al., 2019). Note that we see greater amounts of variance for the validation and test sets than the train set, as is expected given their smaller size of 422 and 511 subjects respectively.

**Figure 4.**
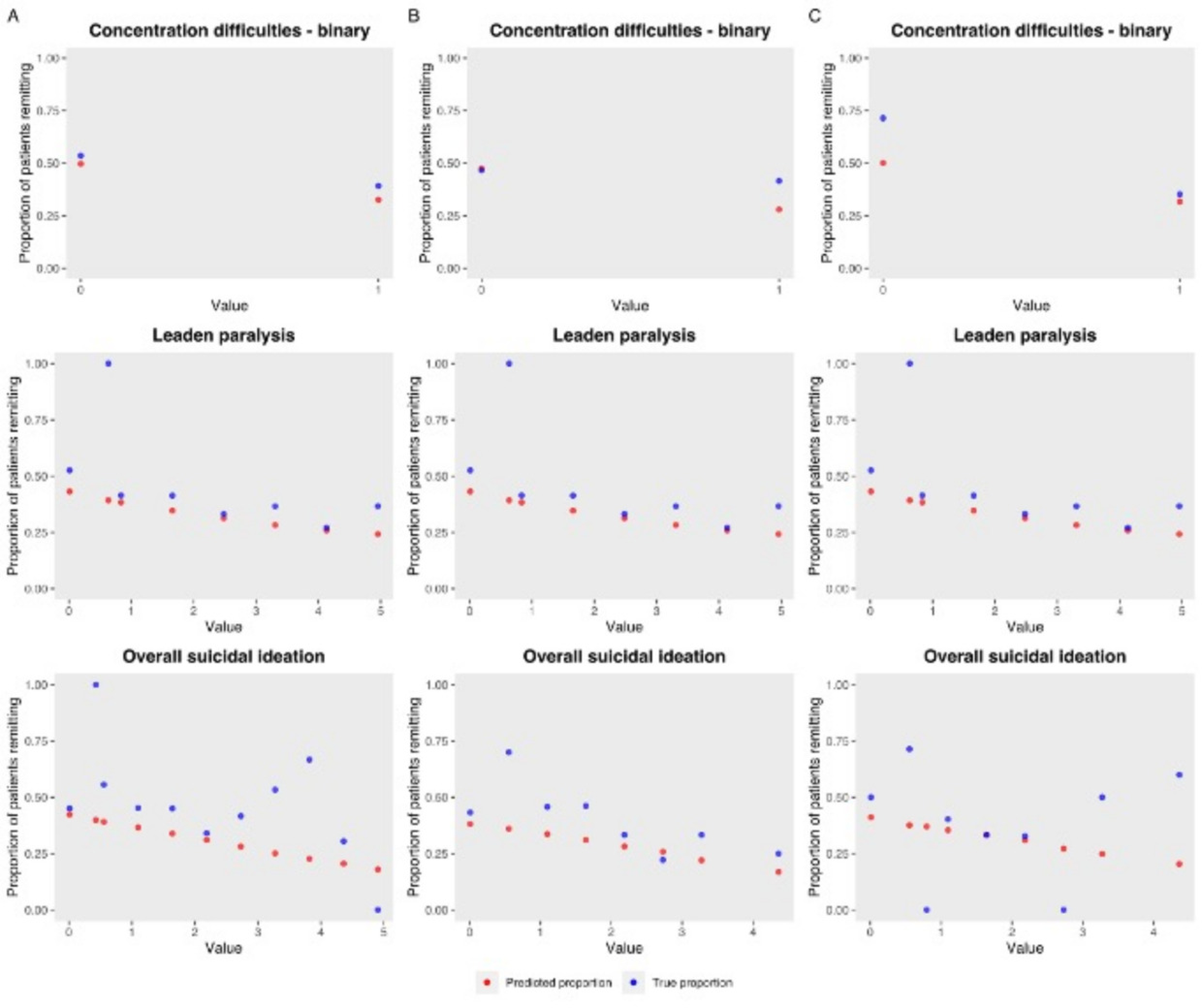
Sensitivity test output examples. These figures show, for each of our A) train (n=4099), B) validation(n=422), and C) test (n=511) sets, the proportion of patients remitting according as the target variable is varied. Of these 3 features, two (recent suicidal ideation, leaden paralysis) possess categorical values, while one (concentration) is an example of a binary feature. The number of data points reflect the possible values that were created during the transformation and standardization process - in some cases values were created between whole numbers in order to best represent partially overlapping scales. Blue dots represent the observed proportion of patients remitting who truly had those values; red dots represent the predicted probabilities when all patients in the dataset have the feature set to the given value. Note that in smaller datasets like in validation or train, where very few patients may naturally have a given value, larger variance occurs in the observed values.

### Bias Testing

Figure 5 provides the results of our post-hoc bias tests examining the difference in the predicted versus the observed (real) probability for each of the subgroups. No group had an under-prediction of remission rate, compared to the true rate, of more than 5%, indicating that the model did not learn to amplify biases for any one group. In supplementary Tables 2-4, the basic statistical metrics used to analyze our subgroup data (race, sex, age) are listed for each of the respective sets (train, validation, test); reasonable metrics are preserved for each subgroup, except when the n for that subgroup is small.

**Figure 5.**
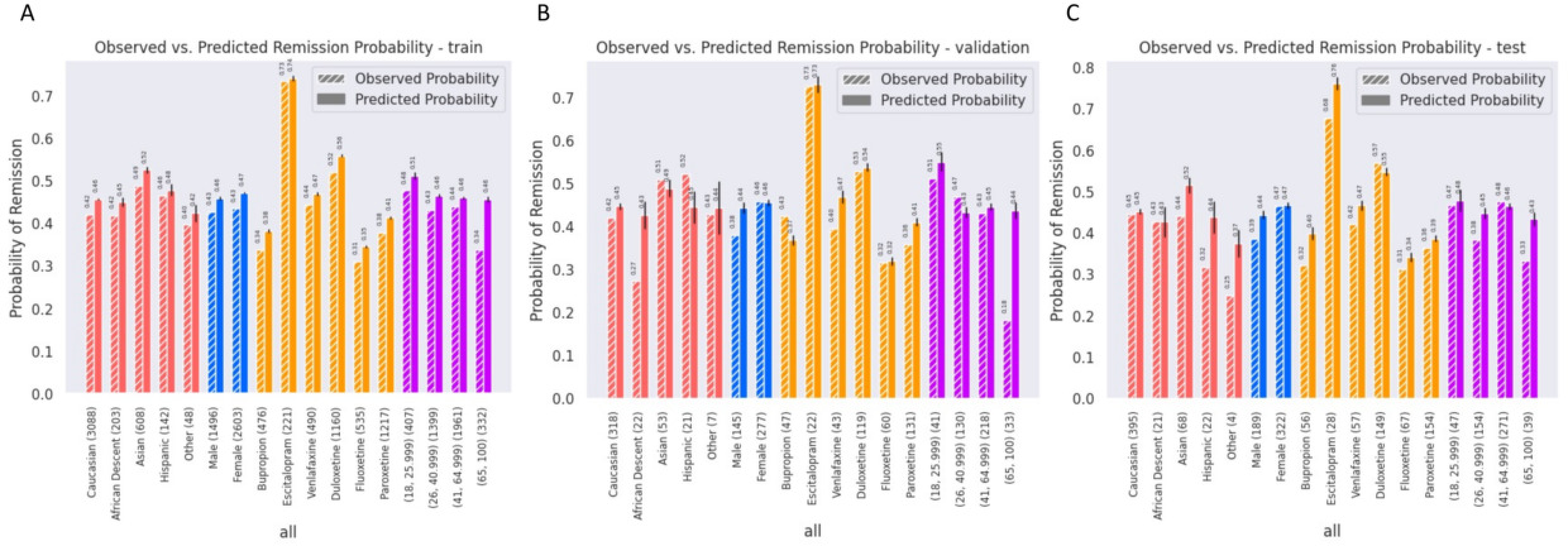
Report on bias testing. This figure shows both the observed and mean predicted probability, including the standard error, of patients remitting based on their race (Caucasian, African Descent, Asian, Hispanic, Other), sex (Male, Female), antidepressant treatments (Bupropion, Escitalopram, Venlafaxine, Fluoxetine, Paroxetine), and age groups (18-25.9; 26-40.9; 41-64.9; 65-130.9) in the a) Train, b) Validation, c) Test sets. The error bars represent the standard error for the predicted probability.

## Discussion

In this study, we present a full pipeline from data preprocessing to model validation that was harnessed to create the first ever differential treatment selection for MDD containing 6 treatment options. We introduce our novel methodology for tackling the heterogeneity of available datasets - particularly the creation of a custom taxonomy and transformation process that expanded upon previous taxonomies which, alone, were unable to capture and integrate the breadth of variables required. We also discuss other challenges, such as the missingness of data the need to transform categorical and binary variables differently. In addition, we demonstrate bias testing through the analysis of model metrics and the comparison of predicted and observed remission rates in key subgroups.

In the literature, there is a general lack of detailed mechanistic understanding regarding factors that impact treatment remission in depression and how they interact with one another (Perlman et al., 2019). Recent work has focused on anxious depression and its correlation with worse prognosis (Gaspersz et al., 2017), and studies have found that certain variables such as psychic anxiety are more negatively predictive of treatment outcomes (Iniesta et al., 2016). Our model also found psychic anxiety to be an important predictive feature (Table 2). Iniesta et al. (2016) found other predictive variables including questions related to apparent sadness, pessimism, and indecisiveness. Sadness, perhaps due to being a common symptom and therefore unlikely to be predictive of differential treatment response, was not identified as an important feature by our model; however, pessimism – captured as “hopeless outlook of future” – did appear on our list of features, as did difficulties with concentration, which may be linked to indecisiveness (APA 2013, Lauderdale et al., 2021). Sleep is another feature identified as predictive by our model, which has also been found to be predictive in previous work (see Perlman et al., 2019 for a review). More specifically, work has shown that prolonged sleep latency and insomnia, both alone and in combination, are predictive for nonremission (Troxel et al., 2011). Moreover, a sleep profile consisting of reduced REM latency, increased REM density, and poor sleep continuity described in older work, was associated with poorer outcomes following psychotherapy (interpersonal and cognitive behavioral therapy, (Thase et al., 1997; Thase et al., 1996) and pharmacotherapy (fluoxetine or imipramine, (Thase et al., 1997)).

Race was found to be an important predictive feature. Our interpretation of this finding stems from the notion that race is often confounded with a number of important sociodemographic features – not available in the dataset, and that this may be driving the effect. Previous studies, such as the STAR*D study, have concluded that black individuals had lower remission rates than white individuals (Panaite et al., 2019) and Hispanic individuals were somewhere in between; crucially, this was before adjusting for social factors and the statistical significance was lost following the adjustments, although Black individuals did still have a lower remission rate (Lesser et al., 2007). A follow-up study argued for the role of genetic ancestry – not race – as accounting for much of the residual disparity even after accounting for socioeconomic and baseline clinical factors (Murphy et al., 2013). As such, clinicians interpreting the results of models trained on datasets such as this one should consider not just race, but other social determinants of health often confounded with race when treating patients.

Sex was also found to be an important feature. This is despite the fact that there is no clear consensus regarding sex-related differences in remission with antidepressant treatment (Sramek et al., 2016). There does seem to be some evidence that serotonergic antidepressants yield better responses in females than males due to the modulating role of estrogen (Berlanga & Flores-Ramos, 2006). What is most likely, however, is that sex is interacting with other features while the prediction is made, as noted for sleep above. These complex, non-linear interactions can be a challenge to interpret, though some headway can be made using classical techniques, though this would be out of scope for this paper (see Mehltretter et al., 2020).

The reader may note that, at the population level, clear trends emerge in our data with respect to the ranking of antidepressants in terms of their predicted effectiveness. These predictions reflect the underlying data (see Figure 5) and are indeed reminiscent of the order of the ranking of treatment efficacy in two large meta-analyses (Cipriani et al., 2009; Cipriani et al., 2018). At the individual patient level, however, the ranking of all treatments changes and, crucially, the remission rate predictions for each treatment varies. The fundamental clinical utility of the model lies in this variation. However, the good performance of some drugs (e.g. escitalopram) may positively skew the predicted benefit result at the population level (this was despite the fact that most trials included here, including escitalopram, had similar inclusion and exclusion criteria). As such, clinical trials are needed to assess the real-world impact of the model on treatment outcomes.

There has been an encouraging recent effort to standardize the assessment instruments used in both clinical practice and clinical studies-for example, the use of the PHQ-9, GAD-7, and WHODAS 2.0 (Waheed et al., 2024). While these questionnaires were not available in the majority of the datasets we had access to for this study, their inclusion in future research should facilitate the development of the next generation of predictive models and simplify pipelines required for their development. At the same time, efforts to standardize assessments may also be informed by work such as that which we present here. For example, the PHQ-9 has a single item covering both reduced and increased appetite, whereas our model identified reduced appetite as a predictive symptom; as such, future work on harmonized instruments might benefit from including more precise items but only in cases where these have been shown to be predictive.

Compared to imaging or genetic-based predictive tools, questionnaire-based assessments may be more easily integrated into clinical practice and may provide results with greater speed as they can be generated as soon as the patient and clinician respond to any required items. However, there can still be significant barriers to implementing questionnaire-based assessments and predictive tools into clinical practice. These barriers can include assessments that are not designed to fit into the clinical workflow, or which are too time-consuming for clinicians or patients to complete, or which are done in an unwieldly manner (e.g., on paper and then uploaded to a computer database, or through a poorly designed computer program). In previous work, we have described a participatory iterative design process in which clinicians and patients are engaged in the development and validation of a computer-based decision support system (Benrimoh et al., 2021; Popescu et al., 2021; Golden et al., 2024). The platform was designed to be used rapidly by clinicians within a clinical appointment, and by patients at home via mobile application with reminders to complete assessments. With this computerized system, both patients and clinicians were shown to use the platform in a consistent manner over a 12-week follow-up period (Popescu et al., 2021)

Once a model has been trained and tested in the manner we describe in this article, it is important to consider how it could then be implemented into clinical practice. The first step would be to define the process for collecting data from future patients. The feature selection performed by the pipeline we describe defines the features that must be collected from patients in order to generate predictions. Once the features are known, a representative question could then be chosen from among the existing validated items used to create the feature. This could be done based on selecting the most common question in the dataset from among those making up the feature, or based on the question whose distribution best matches that of the final feature. The questions selected in this manner could then be administered to patients via a computer interface, and the responses to these questions would then be fed through the pipeline we describe in order to generate data consistent with the features in the model training data. This data could then be used by the model to generate predictions, which would take the format of remission probabilities for each treatment the model was trained on. These remission probabilities would then be presented to clinicians using a computerized decision support platform, such as the one described in (Benrimoh et al., 2021; Popescu et al., 2021), as one more piece of clinical information which could be used to, in collaboration with the patient, make a treatment decision. Indeed, we have proceeded to do this in a clinical trial (NCT04655924) whose results will be reported separately.

Our study has a number of limitations. Firstly, several features had to be discarded because they did not meet the sample size limitations. It remains possible that these features may have predictive power; however, they could not be included in our model given that questions must have a representative population for each treatment. Furthermore, the results of individual studies are not representative of populations that are not included in the data. The use of clinical trial data in this case results in a dataset with a number of exclusion criteria, limiting generalizability. Common exclusion criteria for the included studies are psychiatric comorbidities, many of which, such as personality and mood disorders for instance, are predictors of treatment response. Consequently, the results may not be generalizable to certain patient groups with more complex courses of illness or with treatment resistant depression. There are also some concerns with basic demographic information that was included. Notably, race and ethnicity categories are inconsistent across studies. For instance, some studies have a specific category for Native Americans whereas others may consider this population as part of the ‘Other’ category. Nevertheless, our data did have a wide spread of ages and considerable representation of different ethnicities, though future work needs to include more diverse populations. Moreover, some studies are missing important sociodemographic data such as level of education and socioeconomic status, which are both key variables in predicting remission (Carter et al., 2012). Additionally, the model does not include all possible treatments due to the constraints of the data available; a notable example is mirtazapine. Finally, it is worth noting that our taxonomization process depended on subjective judgements of raters and the larger group; despite the fact that the merged data was validated in the method described above, this initial reliance on a qualitative process does introduce the possibility for biases or errors (see Supplementary Methods for details on how this process was handled, see Supplemental Material for a discussion on bias).

In conclusion, we present a complete pipeline used to produce the first ever differential treatment benefit prediction model for MDD containing 6 first-line medication options Mental health data taken from different studies is remarkably heterogeneous, making attempts at merging datasets in order to facilitate the generation of a differential treatment benefit prediction model inherently difficult. To our knowledge, we are the first to create a detailed pipeline for the merging and transformation of heterogeneous clinical trial datasets variables in order to facilitate the generation of a treatment benefit prediction model. We hope that other researchers in the field of psychiatry and beyond can use this generalizable framework to harness the utility of highly variable yet crucial datasets.

## Acknowledgements

We would like to thank GlaxoSmithKline and Eli Lilly for providing the de-identified individual patient raw data and information for the clinical trials.

## Data availability

All GlaxoSmithKline and Eli Lilly data was obtained via the Clinical Study Data Request (CSDR) platform - we do not own any of the clinical trial data. Studies with the following IDs were utilized in this paper: gsk_29060_128, gsk_29060_115, US-HMCR, gsk_ak130939, gsk-WXL101497, gsk_29060_874, lilly_F1J-MC-HMCQ, lily_F1J-AA-HMCV, gsk_29060_810, gsk_AK1113351, lilly_FJ1-MC-HMAYa, lilly_FJ1-MC-HMAYb, lilly_FJ1-MC-HMATA, lilly_FJ1-MC-HMATb, lilly_FIJ-MC-HMAQb, lilly_F1J-MC-HMBV, lilly_FJ1-MC-HMAQa. All information about the model, including model weights and scaling factors can be found at the following github link: https://github.com/Aifred-Health/pharma_research_model

## Author contributions

DB, JM, RF, CA, KP, and GT conceived of and designed the study. KP, CP, JFT, JW, CR, GG, DB and JM performed the pre-processing and transformation JM, CA, RF, and DB performed the machine learning analyses and validation. GT supervised the study. All authors contributed to the writing and/or editing of the manuscript. KP, JM, and DB contributed equally to this study.

## References

American Psychiatric Association. (2013). DSM-5 diagnostic classification. Diagnostic and statistical manual of mental disorders (10th ed.).

Benrimoh, D., Fratila, R., Israel, S., Perlman, K., Mirchi, N., Desai, S., Rosenfeld, A., Knappe, S., Behrmann, J., Rollins, C., & You, R. P. (2018). Aifred health, a deep learning powered clinical decision support system for mental health. In The NIPS ’17 Competition: Building Intelligent Systems (pp. 251–287). Spinger International Publishing.

Benrimoh, D., Tanguay-Sela, M., Perlman, K., Israel, S., Mehltretter, J., Armstrong, C., Fratila, R., Parikh, S. V., Karp, J. F., Heller, K., Vahia, I. V., Blumberger, D. M., Karama, S., Vigod, S. N., Myhr, G., Martins, R., Rollins, C., Popescu, C., Lundrigan, E., Snook, E., … Margolese, H. C. (2021). Using a simulation centre to evaluate preliminary acceptability and impact of an artificial intelligence-powered clinical decision support system for depression treatment on the physician-patient interaction. BJPsych open, 7(1), e22. 10.1192/bjo.2020.127

Berlanga, C., & Flores-Ramos, M. (2006). Different gender response to serotonergic and noradrenergic antidepressants. A comparative study of the efficacy of citalopram and reboxetine. Journal of Affective Disorders, 95(1–3), 119–123. 10.1016/j.jad.2006.04.029

Blanken, T. F., Borsboom, D., Penninx, B. W., & Van Someren, E. J. (2020). Network outcome analysis identifies difficulty initiating sleep as a primary target for prevention of depression: A 6-year prospective study. Sleep, 43(5), zsz288. 10.1093/sleep/zsz288

Borisov, V., Haug, J., & Kasneci, G. (2019). CancelOut: A Layer for Feature Selection in Deep Neural Networks. In I. V. Tetko, V. Kůrková, P. Karpov, & F. Theis (Eds.), Artificial Neural Networks and Machine Learning – ICANN 2019: Deep Learning (Vol. 11728, pp. 72–83). Springer International Publishing. 10.1007/978-3-030-30484-3_6

Carter, G. C., Cantrell, R. A., Victoria Zarotsky, Haynes, V. S., Phillips, G., Alatorre, C. I., Goetz, I., Paczkowski, R., & Marangell, L. B. (2012). COMPREHENSIVE REVIEW OF FACTORS IMPLICATED IN THE HETEROGENEITY OF RESPONSE IN DEPRESSION: Review: Heterogeneity in Depression. Depression and Anxiety, 29(4), 340–354. 10.1002/da.21918

Caruana, R., Lawrence, S., & Giles, C. L. (2001). Overfitting in Neural Nets: Backpropagation, Conjugate Gradient, and Early Stopping. Neural Information Processing Systems. 402– 408.

Celi, L. A., Cellini, J., Charpignon, M.-L., Dee, E. C., Dernoncourt, F., Eber, R., Mitchell, W. G., Moukheiber, L., Schirmer, J., Situ, J., Paguio, J., Park, J., Wawira, J. G., Yao, S., & for MIT Critical Data. (2022). Sources of bias in artificial intelligence that perpetuate healthcare disparities—A global review. PLOS Digital Health, 1(3), e0000022. 10.1371/journal.pdig.0000022

Chen, R., Stewart, W. F., Sun, J., Ng, K., & Yan, X. (2019). Recurrent Neural Networks for Early Detection of Heart Failure From Longitudinal Electronic Health Record Data: Implications for Temporal Modeling With Respect to Time Before Diagnosis, Data Density, Data Quantity, and Data Type. Circulation: Cardiovascular Quality and Outcomes, 12(10), e005114. 10.1161/CIRCOUTCOMES.118.005114

Cipriani, A., Furukawa, T. A., Salanti, G., Chaimani, A., Atkinson, L. Z., Ogawa, Y., Leucht, S., Ruhe, H. G., Turner, E. H., Higgins, J. P. T., Egger, M., Takeshima, N., Hayasaka, Y., Imai, H., Shinohara, K., Tajika, A., Ioannidis, J. P. A., & Geddes, J. R. (2018). Comparative efficacy and acceptability of 21 antidepressant drugs for the acute treatment of adults with major depressive disorder: A systematic review and network meta-analysis. The Lancet, 391(10128), 1357–1366. 10.1016/S0140-6736(17)32802-7

Clevert, D.-A., Unterthiner, T., & Hochreiter, S. (2015). Fast and Accurate Deep Network Learning by Exponential Linear Units (ELUs). 10.48550/ARXIV.1511.07289

Durstewitz, D., Koppe, G., & Meyer-Lindenberg, A. (2019). Deep neural networks in psychiatry. Molecular Psychiatry, 24(11), 1583–1598. 10.1038/s41380-019-0365-9

Fenton, C., & McLoughlin, D. M. (2021). Usefulness of Hamilton rating scale for depression subset scales and full versions for electroconvulsive therapy. PLOS ONE, 16(11), e0259861. 10.1371/journal.pone.0259861

Gaspersz, R., Lamers, F., Kent, J. M., Beekman, A. T. F., Smit, J. H., van Hemert, A. M., Schoevers, R. A., & Penninx, B. W. J. H. (2017). Longitudinal Predictive Validity of the DSM-5 Anxious Distress Specifier for Clinical Outcomes in a Large Cohort of Patients With Major Depressive Disorder. The Journal of Clinical Psychiatry, 78(02), 207–213. 10.4088/JCP.15m10221

Gokcay, D., Eken, A., & Baltaci, S. (2019). Binary Classification Using Neural and Clinical Features: An Application in Fibromyalgia With Likelihood-Based Decision Level Fusion. IEEE Journal of Biomedical and Health Informatics, 23(4), 1490–1498. 10.1109/JBHI.2018.2844300

Golden, G., Popescu, C., Israel, S., Perlman, K., Armstrong, C., Fratila, R., Tanguay-Sela, M., & Benrimoh, D. (2024). Applying artificial intelligence to clinical decision support in mental health: What have we learned? Health Policy and Technology, 100844. 10.1016/j.hlpt.2024.100844

Greden, J. F., Parikh, S. V., Rothschild, A. J., Thase, M. E., Dunlop, B. W., DeBattista, C., Conway, C. R., Forester, B. P., Mondimore, F. M., Shelton, R. C., Macaluso, M., Li, J., Brown, K., Gilbert, A., Burns, L., Jablonski, M. R., & Dechairo, B. (2019). Impact of pharmacogenomics on clinical outcomes in major depressive disorder in the GUIDED trial: A large, patient- and rater-blinded, randomized, controlled study. Journal of Psychiatric Research, 111, 59–67. 10.1016/j.jpsychires.2019.01.003

Greenberg, P. E., Fournier, A.-A., Sisitsky, T., Pike, C. T., & Kessler, R. C. (2015). The Economic Burden of Adults With Major Depressive Disorder in the United States (2005 and 2010). The Journal of Clinical Psychiatry, 76(02), 155–162. 10.4088/JCP.14m09298

Hyde, J. S., & Mezulis, A. H. (2020). Gender Differences in Depression: Biological, Affective, Cognitive, and Sociocultural Factors. Harvard Review of Psychiatry, 28(1), 4–13. 10.1097/HRP.0000000000000230

Iniesta, R., Malki, K., Maier, W., Rietschel, M., Mors, O., Hauser, J., Henigsberg, N., Dernovsek, M. Z., Souery, D., Stahl, D., Dobson, R., Aitchison, K. J., Farmer, A., Lewis, C. M., McGuffin, P., & Uher, R. (2016). Combining clinical variables to optimize prediction of antidepressant treatment outcomes. Journal of Psychiatric Research, 78, 94–102. 10.1016/j.jpsychires.2016.03.016

Jin Huang, & Ling, C. X. (2005). Using AUC and accuracy in evaluating learning algorithms. IEEE Transactions on Knowledge and Data Engineering, 17(3), 299–310. 10.1109/TKDE.2005.50

Kapelner, A., Bleich, J., Levine, A., Cohen, Z. D., DeRubeis, R. J., & Berk, R. (2014). Evaluating the Effectiveness of Personalized Medicine with Software. 10.48550/ARXIV.1404.7844

Kennedy, S. H., Lam, R. W., McIntyre, R. S., Tourjman, S. V., Bhat, V., Blier, P., Hasnain, M., Jollant, F., Levitt, A. J., MacQueen, G. M., McInerney, S. J., McIntosh, D., Milev, R. V., Müller, D. J., Parikh, S. V., Pearson, N. L., Ravindran, A. V., Uher, R., & the CANMAT Depression Work Group. (2016). Canadian Network for Mood and Anxiety Treatments (CANMAT) 2016 Clinical Guidelines for the Management of Adults with Major Depressive Disorder: Section 3. Pharmacological Treatments. The Canadian Journal of Psychiatry, 61(9), 540–560. 10.1177/0706743716659417

Kolen, M. J., & Brennan, R. L. (2014). Test Equating, Scaling, and Linking. Springer New York. 10.1007/978-1-4939-0317-7

Lederer, J. (2021). Activation Functions in Artificial Neural Networks: A Systematic Overview. 10.48550/ARXIV.2101.09957

Lesser, I. M., Castro, D. B., Gaynes, B. N., Gonzalez, J., Rush, A. J., Alpert, J. E., Trivedi, M., Luther, J. F., & Wisniewski, S. R. (2007). Ethnicity/Race and Outcome in the Treatment of Depression: Results From STAR*D. Medical Care, 45(11), 1043–1051. 10.1097/MLR.0b013e3181271462

Liu, Y., Zhang, N., Bao, G., Huang, Y., Ji, B., Wu, Y., Liu, C., & Li, G. (2019). Predictors of depressive symptoms in college students: A systematic review and meta-analysis of cohort studies. Journal of Affective Disorders, 244, 196–208. 10.1016/j.jad.2018.10.084

Luppa, M., Sikorski, C., Luck, T., Ehreke, L., Konnopka, A., Wiese, B., Weyerer, S., König, H.-H., & Riedel-Heller, S. G. (2012). Age- and gender-specific prevalence of depression in latest-life – Systematic review and meta-analysis. Journal of Affective Disorders, 136(3), 212–221. 10.1016/j.jad.2010.11.033

Mehltretter, J., Fratila, R., Benrimoh, D. A., Kapelner, A., Perlman, K., Snook, E., Israel, S., Armstrong, C., Miresco, M., & Turecki, G. (2020). Differential Treatment Benet Prediction for Treatment Selection in Depression: A Deep Learning Analysis of STAR*D and CO-MED Data. Computational Psychiatry, 4(0), 61. 10.1162/cpsy_a_00029

Mehltretter, J., Rollins, C., Benrimoh, D., Fratila, R., Perlman, K., Israel, S., Miresco, M., Wakid, M., & Turecki, G. (2020). Analysis of Features Selected by a Deep Learning Model for Differential Treatment Selection in Depression. Frontiers in Artificial Intelligence, 2, 31. 10.3389/frai.2019.00031

Michelsen C, Jørgensen CC, Heltberg M, Jensen MH, Lucchetti A, Petersen PB, Petersen T, Kehlet H; Center for Fast-track Hip Knee Replacement Collaborative group; Madsen F, Hansen TB, Gromov K, Jakobsen T, Varnum C, Overgaard S, Rathsach M, Hansen L. Machine-learning vs. logistic regression for preoperative prediction of medical morbidity after fast-track hip and knee arthroplasty-a comparative study. BMC Anesthesiol. 2023 Nov 29;23(1):391. doi: 10.1186/s12871-023-02354-z. PMID: 38030979; PMCID: PMC10685559.

Murphy, E., Hou, L., Maher, B. S., Woldehawariat, G., Kassem, L., Akula, N., Laje, G., & McMahon, F. J. (2013). Race, Genetic Ancestry and Response to Antidepressant Treatment for Major Depression. Neuropsychopharmacology, 38(13), 2598–2606. 10.1038/npp.2013.166

Ni, D., Leonard, J. D., Guin, A., & Feng, C. (2005). Multiple Imputation Scheme for Overcoming the Missing Values and Variability Issues in ITS Data. Journal of Transportation Engineering, 131(12), 931–938. 10.1061/(ASCE)0733-947X(2005)131:12(931)

Nutt, D., Wilson, S., & Paterson, L. (2008). Sleep disorders as core symptoms of depression. Dialogues in Clinical Neuroscience, 10(3), 329–336. 10.31887/DCNS.2008.10.3/dnutt

Panaite, V., Bowersox, N. W., Zivin, K., Ganoczy, D., Kim, H. M., & Pfeiffer, P. N. (2019). Individual and neighborhood characteristics as predictors of depression symptom response. Health Services Research, 54(3), 586–591. 10.1111/1475-6773.13127

Pedregosa, F., Varoquaux, G., Gramfort, A., Michel, V., Thirion, B., Grisel, O., Blondel, M., Müller, A., Nothman, J., Louppe, G., Prettenhofer, P., Weiss, R., Dubourg, V., Vanderplas, J., Passos, A., Cournapeau, D., Brucher, M., Perrot, M., & Duchesnay, É. (2012). Scikit-learn: Machine Learning in Python. 10.48550/ARXIV.1201.0490

Perlman, K., Benrimoh, D., Israel, S., Rollins, C., Brown, E., Tunteng, J.-F., You, R., You, E., Tanguay-Sela, M., Snook, E., Miresco, M., & Berlim, M. T. (2019). A systematic meta-review of predictors of antidepressant treatment outcome in major depressive disorder. Journal of Affective Disorders, 243, 503–515. 10.1016/j.jad.2018.09.067

Popescu, C., Golden, G., Benrimoh, D., Tanguay-Sela, M., Slowey, D., Lundrigan, E., Williams, J., Desormeau, B., Kardani, D., Perez, T., Rollins, C., Israel, S., Perlman, K., Armstrong, C., Baxter, J., Whitmore, K., Fradette, M. J., Felcarek-Hope, K., Soufi, G., Fratila, R., … Turecki, G. (2021). Evaluating the Clinical Feasibility of an Artificial Intelligence-Powered, Web-Based Clinical Decision Support System for the Treatment of Depression in Adults: Longitudinal Feasibility Study. JMIR formative research, 5(10), e31862. 10.2196/31862

Puzhko, S., Aboushawareb, S. A. E., Kudrina, I., Schuster, T., Barnett, T. A., Renoux, C., & Bartlett, G. (2020). Excess body weight as a predictor of response to treatment with antidepressants in patients with depressive disorder. Journal of Affective Disorders, 267, 153–170. 10.1016/j.jad.2020.01.113

Raskutti, G., Wainwright, M. J., & Yu, B. (2011). Early stopping for non-parametric regression: An optimal data-dependent stopping rule. 2011 49th Annual Allerton Conference on Communication, Control, and Computing (Allerton), 1318–1325. 10.1109/Allerton.2011.6120320

Ruggero, C. J., Kotov, R., Hopwood, C. J., First, M., Clark, L. A., Skodol, A. E., Mullins-Sweatt, S. N., Patrick, C. J., Bach, B., Cicero, D. C., Docherty, A., Simms, L. J., Bagby, R. M., Krueger, R. F., Callahan, J. L., Chmielewski, M., Conway, C. C., De Clercq, B., Dornbach-Bender, A., Eaton, N. R., … Zimmermann, J. (2019). Integrating the Hierarchical Taxonomy of Psychopathology (HiTOP) into clinical practice. Journal of consulting and clinical psychology, 87(12), 1069–1084. 10.1037/ccp0000452

Salk, R. H., Hyde, J. S., & Abramson, L. Y. (2017). Gender differences in depression in representative national samples: Meta-analyses of diagnoses and symptoms. Psychological Bulletin, 143(8), 783–822. 10.1037/bul0000102

Seyyed-Kalantari, L., Zhang, H., McDermott, M. B. A., Chen, I. Y., & Ghassemi, M. (2021). Underdiagnosis bias of artificial intelligence algorithms applied to chest radiographs in under-served patient populations. Nature Medicine, 27(12), 2176–2182. 10.1038/s41591-021-01595-0

Snoek, J., Larochelle, H., & Adams, R. P. (2012). Practical Bayesian Optimization of Machine Learning Algorithms. 10.48550/ARXIV.1206.2944

Squarcina, L., Villa, F. M., Nobile, M., Grisan, E., & Brambilla, P. (2021). Deep learning for the prediction of treatment response in depression. Journal of Affective Disorders, 281, 618– 622. 10.1016/j.jad.2020.11.104

Sramek, J. J., Murphy, M. F., & Cutler, N. R. (2016). Sex differences in the psychopharmacological treatment of depression. Dialogues in Clinical Neuroscience, 18(4), 447–457. 10.31887/DCNS.2016.18.4/ncutler

Sung, S. C., Wisniewski, S. R., Luther, J. F., Trivedi, M. H., & Rush, A. J. (2015). Pre-treatment insomnia as a predictor of single and combination antidepressant outcomes: A CO-MED report. Journal of Affective Disorders, 174, 157–164. 10.1016/j.jad.2014.11.026

Tanguay-Sela, M., Benrimoh, D., Popescu, C., Perez, T., Rollins, C., Snook, E., Lundrigan, E., Armstrong, C., Perlman, K., Fratila, R., Mehltretter, J., Israel, S., Champagne, M., Williams, J., Simard, J., Parikh, S. V., Karp, J. F., Heller, K., Linnaranta, O., … Margolese, H. C. (2022). Evaluating the perceived utility of an artificial intelligence-powered clinical decision support system for depression treatment using a simulation center. Psychiatry Research, 308, 114336. 10.1016/j.psychres.2021.114336

Tharwat, A. (2020). Classification assessment methods. Applied Computing and Informatics. 17: 168–192.

Vink, D., Aartsen, M. J., & Schoevers, R. A. (2008). Risk factors for anxiety and depression in the elderly: A review. Journal of Affective Disorders, 106(1–2), 29–44. 10.1016/j.jad.2007.06.005

Waheed, A., Afridi, A. K., Rana, M., Arif, M., Barrera, T., Patel, F., Khan, M. N., & Azhar, E. (2024). Knowledge and Behavior of Primary Care Physicians Regarding Utilization of Standardized Tools in Screening and Assessment of Anxiety, Depression, and Mood Disorders at a Large Integrated Health System. Journal of primary care & community health, 15, 21501319231224711. 10.1177/21501319231224711

Waszczuk, M. A., Kotov, R., Ruggero, C., Gamez, W., & Watson, D. (2017). Hierarchical structure of emotional disorders: From individual symptoms to the spectrum. Journal of Abnormal Psychology, 126(5), 613–634. 10.1037/abn0000264

World Health Organization. (2017). Depression and Other Common Mental Disorders: Global Health Estimates. https://apps.who.int/iris/bitstream/handle/10665/254610/W?sequence=1

Yu, K.-H., & Kohane, I. S. (2019). Framing the challenges of artificial intelligence in medicine. BMJ Quality & Safety, 28(3), 238–241. 10.1136/bmjqs-2018-008551

