## Supplementary Material for "Development of a differential treatment selection model for depression on consolidated and transformed clinical trial datasets"

**SUPPLEMENTARY INFORMATION**

**Supplemental methods**

*Generation of train, validation, and test sets*

When producing our train, validation, and test sets, to ensure they were not skewed by any single outcome or treatment each split was stratified based on the binary remission and the treatment variable. Since we are stratifying on more than one variable and producing three separate sets, the best approach was to sequentially split the data. Using scikit-learn’s *train_test_split function* we took our combined data and created the test set by performing a 90-10 stratified split (Pedregosa et al., 2012). The remaining 90% of the data was then split again to create the 10% validation and test sets. These two splits resulted in our 80-10-10 data set split.

A common machine learning problem is the missingness that exists within the data. To handle this problem, we first established how much missingness is too much, and we did so by visualizing where the missingness existed. To do this we tested three null-value thresholds 50%, 55%, and 65% by removing all features that had a null percentage above the given threshold. For each of these thresholds we assessed how the missingness was distributed across study, drug, and subject. We wanted to ensure that no single study, drug, or subject had a large amount of missing data. While looking at the raw percentage of missing data we also used pie charts, heatmaps and histograms to analyze the missingness across those three key variables. Based on the results of the analyses using the 50%, 55% and 60% thresholds we determined through consensus that 50% was the optimal threshold to ensure that no study, drug or group of subjects had excessive missingness. We used a heatmap plot to analyze the percentage of missing values for each variable in each study and each variable in each drug subgroup. The heatmap showed us if specific studies or drugs were missing many values for specific variables or if a variable was missing many values across many studies or drugs. If a study was missing data for many variables or a variable was missing across many studies or drugs, then we removed the study or the variable. We used a bar plot to assess the missingness of sociodemographic variables across each study and removed studies where much of the demographic data was missing. Decisions were made by group consensus. Lastly, we used a bar plot to analyze missingness from an individual patient level. For each patient, we calculated the percentage of missing values, removing subjects that were missing more than 50% of the values.

For imputing missing data we used multiple imputation by chained equations (MICE) provided by the package Autoimpute (<https://autoimpute.readthedocs.io/en/latest/>), which was built based on the R *mice* function. Given that our neural network architecture did not support the concurrent use of many MICE imputed datasets, we averaged the results from the iterations (Chen et al., 2019; Gokcay et al., 2019; Ni et al., 2005). The package requires various user-specified variables that allow for optimizing the imputation results. For our imputation purposes, we specified the number of imputation iterations, and the strategies and predictors to use for a given variable to be imputed. We adjusted these parameters based on our visualization of missingness for each variable and the evaluation of the imputation results. We evaluated imputation results using a histogram that displays the distribution of an imputed variable before and after imputation. Both distributions are displayed on the same plot which allows us to easily visualize any differences in distribution caused by imputation. If a noticeable difference in distribution occurred for a given variable, we specifically analyzed that variable and tested adjustments to the imputation parameters, variable strategies or variable predictors.

For categorical variables, we used logistic regression from the scikit-learn package. For continuous variables, we use the least-squares strategies or predictive mean matching, both of which also rely on scikit-learn models. Least-squares regression was our default for continuous variables; however, in situations where a continuous variable was skewed toward one class, we used predictive mean matching as the strategy. For the variable predictors, we first restricted possible predictors to be those with high outflux and low influx values. Then from an individual feature perspective, we removed predictor candidates in two cases. The first being candidate predictors that had high multicollinearity with the to-be imputed variable. The second being candidate predictors that had a low intersection of non-null values with the to-be imputed variable.

We fit the imputer with 10 iterations using the train data and used it to impute the missing data on the train, test, and validation data. By training our imputer only on the training set we ensure that no imputations are made based on any information learned from our test or validation set. To validate the imputation results we performed visualized assessments through the use of scatter plots, distribution plots, and boxplots. These assessments allowed us to verify that our imputation did not produce a meaningful difference in the distribution of values or outlier values. Below is an example of the graphic produced after imputation that allows us to confirm there is no significant variation in the distribution after imputation. Continuous variables existed within the dataset which implies the possibility of imputing negative and zero values. A value less than or equal to zero can have a negative impact on neural network training when using certain activation functions. For example, the ReLU activation outputs a value of zero when any negative input is provided (Lederer, 2021). To avoid this type of behaviour from occurring, we adjusted each variable by adding the absolute value of the smallest value available plus a constant 0.01 to the entire variable. For example, if -1 was the smallest value for variable x we added 1.01 to all values of variable x resulting in 0.01 now being the smallest value.

*Bayesian optimization*

A candidate model is prepared by arranging a set of hyperparameters (e.g. number of layers, number of nodes per layer, activation function, learning rate, number of training epochs) to be integrated through by Bayesian Optimization (Snoek et al., 2012). This process leveraged the metrics outlined in methods and supplemental methods section, provided by a 10-fold cross validation, to determine which next set to configure the model with to achieve better performance than the last iteration. Within a training iteration, we make use of Early Stopping which is responsible for checkpointing model states and preserving optimal generalizability of the proposed model by tracking the validation accuracy within a training interval (Caruana et al., 2001; Raskutti et al., 2011). If after a certain number of full training iterations, also optimized (e.g. a patience of 3 epochs) the model is seen to have a steady drop in validation accuracy while the training accuracy continues to improve, this indicates that the model is beginning to overfit on the data and training is stopped (Raskutti et al., 2011). We performed several different experiments using different performance targets for Bayesian optimization in order to search the model space for a model which performed well across all metrics. Crucially, the test set was not used until the final candidate model was chosen.

**Supplementary Results**

*Logistic Regression Comparator Model*

In order to determine if deep learning provided an improvement over more commonly used machine learning approaches, such as logistic regression, we performed a logistic regression trained on the same training set as the deep learning model and tested on the same test set. Importantly, the model utilized the same features that were identified by the deep learning model, and as such it benefitted from the feature selection performed by the neural network. With respect to our primary metric, AUC, the logistic regression model performed worse than the deep learning model, achieving an AUC of 0.62 compared to the deep learning model AUC of 0.7 on the test set. The logistic regression model slightly underperformed the deep learning model in terms of accuracy, achieving an accuracy of 0.65, compared to the deep learning model's accuracy of 0.66 on the test set. This result likely points to the importance of feature selection, which in our neural network architecture is implemented as part of the model training, rather than requiring a separate model. Overall, this analysis suggests that, in line with previous work (Michelsen et al., 2023) that deep learning can outperform other methods, and will likely continue to do so as datasets continue to grow.

**Supplementary discussion on bias**

One key criticism of machine learning models is that they can learn harmful biases from data, include these biases as part of the latent factors influencing their predictions, and perpetuate or amplify these biases (Yu & Kohane, 2019). For example, a study by Seyyed-Kalantari et al., (2021) investigated underdiagnosis in chest X-ray prediction models. The model was found to have a high rate of underdiagnosis among patients in subpopulations with limited access to care (Seyyed-Kalantari et al., 2021). Specifically, patients that are under the age of 20, female, African-American, Hispanic, and from lower socioeconomic backgrounds had higher rates of underdiagnosis (Seyyed-Kalantari et al., 2021).  One must differentiate between the amplification of bias, and the reasonable representation of unfortunate but real associations within the data. Let us take minority status as an example. For various reasons including socioeconomic factors, minority groups may have lower rates of remission than non-minority groups (Celi et al., 2022). Given that this pattern is expected to be observed, it would be reasonable to expect that a representative sample would replicate this effect. In the same way, in order to be a valid representation of relationships within the training data (when, crucially, this data is a valid representation of the population on which the model is to be used), a machine learning model trained on a representative dataset should reproduce these associations to a reasonable degree but, in order to be an ethical model, should *not* amplify them (Hall et al., 2022).  It should be noted that in the case where the data used for model training is not representative of the intended use population, then it is unlikely the model will be free of bias when applied to the intended use population. For illustration, an example of bias amplification is gender bias in natural language processing (NLP), a subset of AI focused on human language (Costa-jussa 2019). In automated translations, examples in which the gender used in a translated sentence is associated with the gender stereotype, even if the inputted sentence is gender neutral (Costa-jussa, 2019). Previous research by Zhao et al., has shown that an NLP model trained on datasets from the internet significantly amplified associations between gender and a frequently gendered activity (i.e. cooking) (2017). While gender bias does already exist in society, NLP models often amplify these existing biases (Bansal, 2022).

We performed subgroup analysis in order to examine the effects of these biases. Great effort was made to minimize these harmful biases; however, we are limited by the data itself. For instance, caucasians are overly represented in the data available. Given that we must work within the constraints of our data, our priority was to ensure harmful biases are not multiplied by the model, which generally does not seem to be the case based on our subgroup analyses.

**Supplementary Table 1**

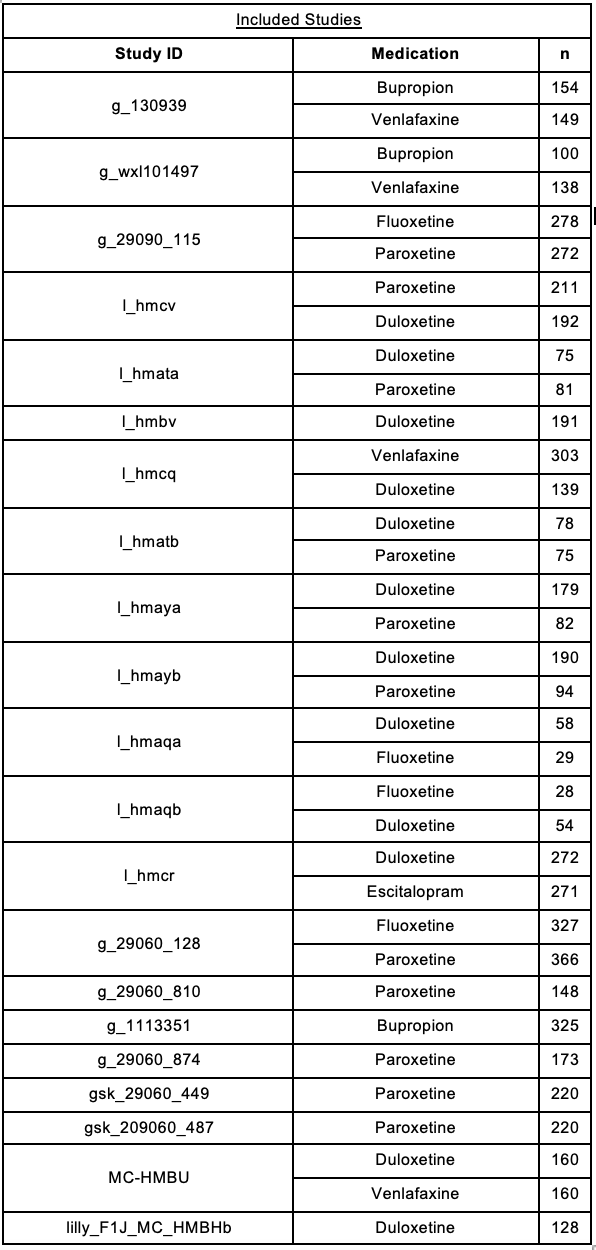

**Supplementary Table 2**

**Glossary**

| **Term** | **Definition** |
| --- | --- |
| Taxonomy | Dimensional classification method. Made up of categories in a tree-like format composed of broader categories that branch into more granular subcategories. Used to categorize questionnaire data in order to facilitate later combining of semantically similar questions. |
| Tree structure (root, branch, leaf) | The taxonomy was created as a tree-like structure. The base categories, which are the most general (e.g., quality of life) are called the roots. All of the subcategories that project off of the main category are called branches (e.g., functional impairment, relationships, mobility). The most granular of all the subcategories (e.g., family, social, romantic) projecting from the main category are called leaves. |
| Standard question | Individual questions from questionnaires administered in the trials, listed with response text and response values. These questions are annotated with the taxonomic system to sort and classify them. |
| Transformed question | Grouping of semantically similar or identical standard questions, whose response values are transformed such that data across this new feature could be “merged” across the whole dataset. |
| Flag | An indication of a specifier to any given question. For example, there is a flag to indicate whether the question is clinician or patient rated and another flag to indicate if questions are referring to past or present time. |
| Equipercentile scaling | An equating method which uses the underlying value percentiles between two variables to convert one scale to be on the same scale as the other variable. |

**Supplemental Table 3**

**Training Set (n = 4099)**

a) Race

| Race | Accuracy | Sensitivity | Specificity | PPV | NPV | F1 | AUC |
| --- | --- | --- | --- | --- | --- | --- | --- |
| Caucasian (3088) | .65 | .50 | .76 | .60 | .67 | .54 | .69 |
| African Descent (203) | .65 | .49 | .76 | .6 | .67 | .54 | .67 |
| Asian (608) | .67 | .70 | .64 | .65 | .69 | .67 | .73 |
| Hispanic (142) | .66 | .56 | .75 | .66 | .66 | .6 | .72 |
| Other (48) | .87 | .42 | .82 | .61 | .68 | .5 | .74 |

b) Sex

| Sex | Accuracy | Sensitivity | Specificity | PPV | NPV | F1 | AUC |
| --- | --- | --- | --- | --- | --- | --- | --- |
| Male (1496) | .66 | .51 | .77 | .62 | .68 | .56 | .70 |
| Female (2603) | .65 | .54 | .73 | .61 | .67 | .57 | .70 |

c) Age

| Age Group | Accuracy | Sensitivity | Specificity | PPV | NPV | F1 | AUC |
| --- | --- | --- | --- | --- | --- | --- | --- |
| (18, 25.9) (407) | .63 | .63 | .64 | .61 | .65 | .62 | .69 |
| (26, 40.9) (1399) | .67 | .55 | .75 | .63 | .69 | .59 | .71 |
| (41, 64.9) (1961) | .65 | .50 | .76 | .63 | .66 | .56 | .71 |
| (65, 130.9) (332) | .61 | .44 | .70 | .43 | .71 | .44 | .62 |

**Supplementary Table 4**

**Validation Set (n = 422)**

a) Race

| Race | Accuracy | Sensitivity | Specificity | PPV | NPV | F1 | AUC |
| --- | --- | --- | --- | --- | --- | --- | --- |
| Caucasian (318) | .65 | .50 | .77 | .61 | .67 | .55 | .66 |
| African Descent (22) | .68 | .33 | .81 | .4 | .76 | .36 | .63 |
| Asian (53) | .6 | .59 | .61 | .61 | .59 | .60 | .56 |
| Hispanic (21) | .66 | .54 | .80 | .75 | .61 | .63 | .64 |
| Other (7) | .85 | .66 | 1.0 | 1.0 | .8 | .8 | .75 |

b) Sex

| Sex | Accuracy | Sensitivity | Specificity | PPV | NPV | F1 | AUC |
| --- | --- | --- | --- | --- | --- | --- | --- |
| Male (145) | .68 | .54 | .76 | .58 | .73 | .56 | .65 |
| Female (277) | .63 | .49 | .75 | .63 | .64 | .55 | .64 |

c) Age

| Age Group | Accuracy | Sensitivity | Specificity | PPV | NPV | F1 | AUC |
| --- | --- | --- | --- | --- | --- | --- | --- |
| (18, 25.9) (41) | .48 | .61 | .34 | .5 | .46 | .55 | .52 |
| (26, 40.9) (130) | .69 | .52 | .84 | .744 | .66 | .61 | .70 |
| (41, 64.9) (218) | .66 | .51 | .79 | .64 | .68 | .57 | .67 |
| (65, 130.9) (33) | .60 | 0.0 | .74 | 0.0 | .76 | 0.0 | .21 |

**Supplementary Table 5**

**Test Set (n = 511)**

a) Race

| Race | Accuracy | Sensitivity | Specificity | PPV | NPV | F1 | AUC |
| --- | --- | --- | --- | --- | --- | --- | --- |
| Caucasian (395) | .65 | .49 | .78 | .65 | .66 | .56 | .71 |
| African Descent (21) | .76 | .55 | .91 | .83 | .73 | .66 | .78 |
| Asian (68) | .60 | .66 | .55 | .54 | .67 | .59 | .68 |
| Hispanic (22) | .68 | .57 | .73 | .5 | .78 | .53 | .67 |
| Other (4) | .75 | 0.0 | 1.0 | 0.0 | .75 | 0.0 | 0.0 |

b) Sex

| Sex | Accuracy | Sensitivity | Specificity | PPV | NPV | F1 | AUC |
| --- | --- | --- | --- | --- | --- | --- | --- |
| Male (189) | .67 | .52 | .77 | .59 | .72 | .55 | .67 |
| Female (322) | .64 | .52 | .75 | .65 | .64 | .57 | .72 |

| Age Group | Accuracy | Sensitivity | Specificity | PPV | NPV | F1 | AUC |
| --- | --- | --- | --- | --- | --- | --- | --- |
| (18, 25.9) (47) | .65 | .59 | .72 | .65 | .66 | .61 | .65 |
| (26, 40.9) (154) | .72 | .55 | .82 | .66 | .75 | .60 | .75 |
| (41, 64.9) (271) | .62 | .50 | .73 | .63 | .61 | .56 | .69 |
| (65, 130.9) (39) | .64 | .38 | .76 | .45 | .71 | .41 | .55 |

**Supplementary Figure 1**
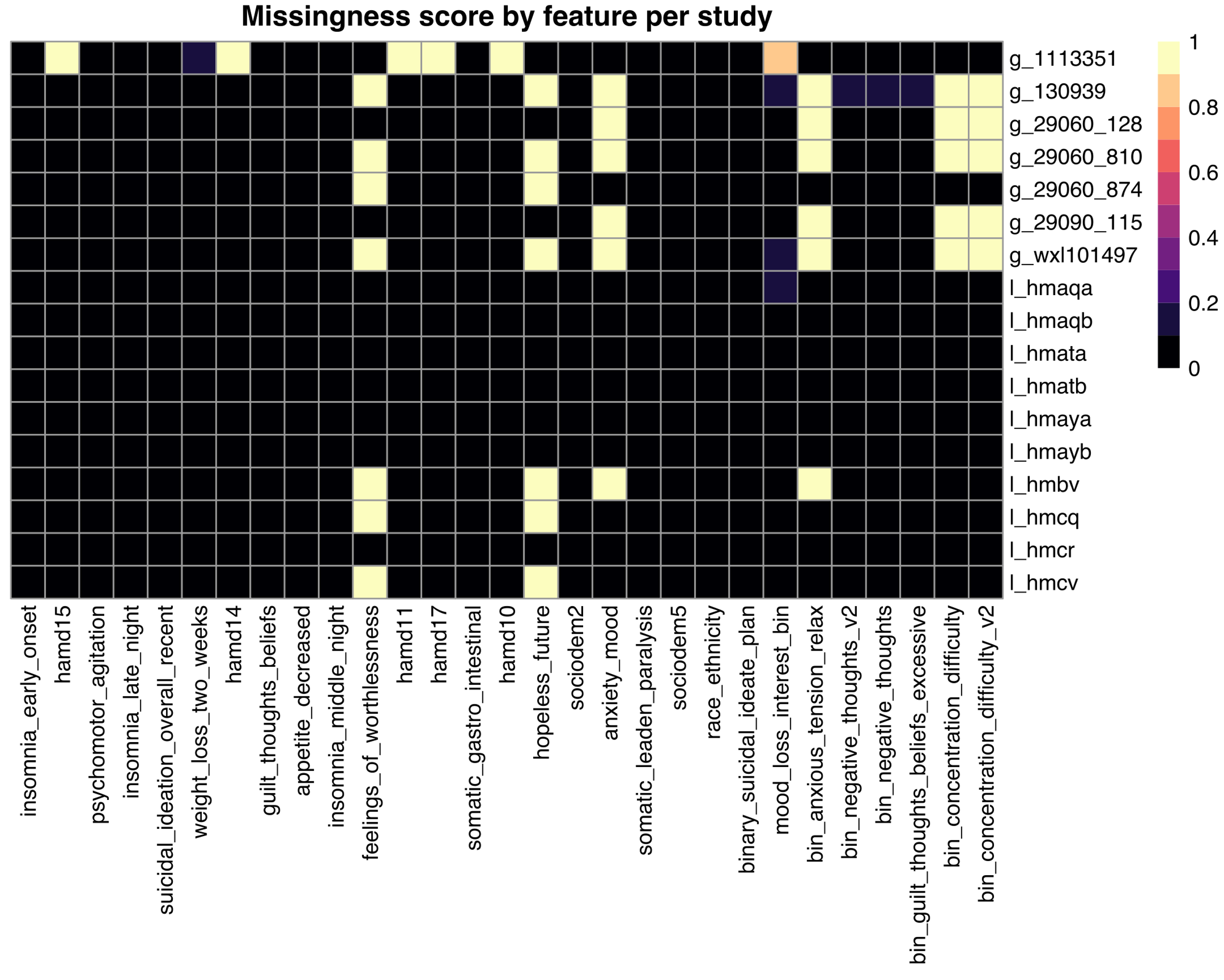

**Missingness score by feature per study**. This heatmap shows the “missingness score” between 0 and 1 as depicted visually by a color bar. A score of 0 indicates no data missingness (i.e., all subjects in this study have data available for that given feature), and a score of 1 indicates complete missingness (i.e., no subjects in that study have data available for that given feature). This heatmap combines the train, validation, and test sets.
